# The inhibition of Macrophage Migration Inhibitory Factor by ISO-1 attenuates trauma-induced multi organ dysfunction in rats

**DOI:** 10.1101/2021.04.28.21255719

**Authors:** Lukas Martin, Nikita Mayur Patel, Noriaki Yamada, Filipe Rodolfo Moreira Borges Oliveira, Lara Stiehler, Elisabeth Zechendorf, Daniel Hinkelmann, Sandra Kraemer, Christian Stoppe, Massimo Collino, Debora Collotta, Gustavo Ferreira Alves, Hanna Pillmann Ramos, Regina Sordi, Ingo Marzi, Borna Relja, Gernot Marx, Christoph Thiemermann

**Affiliations:** Department of Intensive Care and Intermediate Care, University Hospital RWTH Aachen, Aachen, Germany; William Harvey Research Institute, Barts and The London School of Medicine and Dentistry, Queen Mary University of London, London, United Kingdom; Gifu University Graduate School of Medicine, Dept. of Emergency and Disaster Medicine Gifu University Hospital Advanced Critical Care Center; Department of Anesthesiology & Intensive Care Medicine, University Hospital Würzburg, Würzburg, Germany; Department of Drug Science and Technology, University of Turin, Turin, Italy; Department of Pharmacology, Universidade Federal de Santa Catarina, SC, Brazil; Department of Trauma, Hand and Reconstructive Surgery, University Hospital Frankfurt, Goethe University, Frankfurt, Germany; Experimental Radiology, Department of Radiology and Nuclear Medicine, Otto-von-Guericke University, Magdeburg, Germany

**Keywords:** macrophage migratory inhibition factor, ISO-1, hemorrhagic shock, multi organ dysfunction syndrome, trauma

## Abstract

**Background:** Multi organ dysfunction syndrome caused by systemic inflammation after trauma is responsible for a high number of deaths worldwide. The cytokine Migration Inhibitory Factor (MIF) is recognized as a modulator of inflammatory response, however, its role in trauma is unknown. The aim of this study was to investigate (a) the levels of MIF in serum of trauma patients and of rats after hemorrhagic shock, (b) the potential of the MIF tautomerase activity inhibitor ISO-1 to reduce multi organ dysfunction syndrome in an acute and chronic hemorrhagic shock rat model and (c) whether treatment with ISO-1 attenuates NF-κB and NLRP3 activation in hemorrhagic shock.

**Methods:** The serum MIF-levels in trauma patients and rats with hemorrhagic shock were measured by Enzyme-Linked Immunosorbent Assays. Acute and chronic hemorrhagic shock rat models were performed to determine the influence of ISO-1 on multi organ dysfunction syndrome. The activation of NF-κB and NLRP3 pathways were analyzed by western blot.

**Results:** We demonstrated that (a) MIF levels are increased in serum of trauma patients on arrival in the emergency room and in serum of rats after hemorrhagic shock, (b) hemorrhagic shock caused organ damage and low blood pressure (after resuscitation) in rats, while (c) treatment of hemorrhagic shock rats with ISO-1 attenuated organ injury and dysfunction in acute and chronic hemorrhagic shock rat models and (d) decreased the activation of NF-κB and NLRP3 pathways.

**Conclusion:** Our results point to a role of MIF in the pathophysiology of the organ injury and dysfunction caused by trauma/hemorrhage and indicate that MIF tautomerase activity inhibitors may have potential in the therapy of the multi organ dysfunction syndrome after trauma and/or hemorrhage.

## Background

Trauma caused by accidents or violence is the leading cause of death among young people (age 9-49 years) and responsible for over 9 % of all deaths worldwide. According to the World Health Organization, injury is responsible for 6 million deaths per year ^1, 2^. The causes for death as a result of trauma can be divided into three categories: Hemorrhagic shock (HS), brain injury and multi organ dysfunction syndrome (MODS). About 40 % of trauma deaths are related to HS ^3-5^. The numbers of acute deaths decreased in recent years secondary to improved care in the pre-hospital setting ^6^. However, this was associated with an increase in trauma-associated MODS, which is the main cause of death in late post-injury phase ^7, 8^. Post-traumatic MODS is associated with longer stays in the intensive care units (ICU) and is responsible for high numbers of deaths and high costs for the health care system, as there is no specific, preventive treatment ^6, 9, 10^. Organ failure can be categorized in primary and secondary organ failure. Primary organ failure is caused directly by trauma, whereas secondary organ failure is the result of a trauma-induced inflammation and/or organ ischemia ^11^. The systemic inflammation after trauma is triggered by a multitude of factors including poor oxygen supply due to HS and release of damage-associated molecular patterns (DAMPs) caused by tissue injury ^12^. DAMPs work as activators of the immune system and lead to a release of cytokines that can cause severe organ damage ^13^. Furthermore, high cytokine levels are associated with worse prognosis in critically ill patients ^14-16^. The cytokine and enzyme Macrophage Migration Inhibitory Factor (MIF) activates different signaling pathways, including the c-Jun N-terminal kinase pathway and plays a pivotal role in many diseases associated with local or systemic inflammation including sepsis ^17, 18^. Some findings demonstrated that increased MIF levels are associated with poor outcome in patients with sepsis, systemic inflammatory response syndrome and other critical illnesses, whereas other studies highlighted protective effects ^19-21^. The inhibition of MIF’s tautomerase activity by a small molecular weight inhibitor ISO-1 improves outcome in a murine model of sepsis ^22^. ISO-1 inhibits the enzymatic side D-dopachrome tautomerase of MIF, may be of relevance for the pro-inflammatory effects of MIF ^23, 24^. The role of MIF in the pathophysiology of post-traumatic MODS is sparsely known ^25^. Therefore, the aim of this study was to investigate the role of MIF in trauma patients and the effect of ISO-1 on organ failure in rat models of HS.

## Methods

### Study Population and sample collection

A total of 208 patients were included in this study. As described before, the injury severity was evaluated by using the Injury Severity Score (ISS) ^27, 28^. Blood samples from patients with blunt or penetrating trauma with an ISS ≥ 16 were obtained at arrival in emergency room (day 0; A); after 2 days (B); after 5 days (C) and after 7 days (D). The inclusion and exclusion criteria are summarized in Supplemental Table 1. Blood samples were collected in pre-chilled ethylenediaminetetraacetic acid tubes (BD vacutainer, Becton Dickinson Diagnostics, Aalst, Belgium) and kept on ice. Blood was centrifuged at 2000 g for 15 min at 4 °C to separate serum. Sera were stored at ™80 °C for further analysis.

### Experimental Design

Male Wistar rats (for acute model: Charles River Laboratories Ltd., Kent, UK; for chronic model: Universidade Federal de Santa Catarina [UFSC], Brazil) weighing 250-350 g were kept under standard laboratory conditions (12 h light/dark cycle with the temperature maintained at 19–22 °C) and received a chow diet and water *ad libitum*. All animals were allowed to acclimatize to laboratory conditions for at least one week before undergoing any experimentation. (S,R)-3-(4-hydroxyphenyl)-4,5-dihydro-5-isoxazole acetic acid methyl ester (ISO-1) was diluted in 5 % DMSO + 95 % Ringer’s Lactate (vehicle) and rats were treated (i.v. in acute and i.p. in chronic model) upon resuscitation.

### Acute Hemorrhagic Shock Model

The acute hemorrhagic shock model was performed as described previously in this journal ^29, 30^. Specifically, 40 rats were randomized into four groups (n = 10 per group): Sham + vehicle; Sham + ISO-1 (25 mg/kg), HS + vehicle; HS + ISO-1 (25 mg/kg). Detailed description of the acute hemorrhagic shock model and the sample collection as well as a schematic representation of the acute HS model can be found in the supplemental section (Supplemental Figure 1A).

### Chronic Hemorrhagic Shock Model

Thirty rats were randomized into three groups: Sham + vehicle (n = 6); HS + vehicle (n = 12); HS + ISO-1 (25 mg/kg; n = 12). At 24 h post-resuscitation, rats were anesthetized with ketamine-xylazine (100 mg/kg ketamine, 10 mg/kg xylazine i.m.) and samples were collected. Cannulation of the left carotid artery with a polyethylene catheter was performed to measure the MAP and HR; after which up to 5 mL blood was taken via the carotid artery into non-heparinized blood collection tubes. The blood was centrifuged (10000 g for 5 min) to obtain the serum, which was subsequently stored at -80 °C until analysis. Organ collection and measurement of organ injury/dysfunction parameters (Hospital Universitário Professor Polydoro Ernani de São Thiago, Brazil) was performed as described in the acute model. Detailed description as well as a schematic representation of the chronic HS model can be found in the supplemental section (Supplemental Figure 1B).

### MIF enzyme-linked immunosorbent assay

Human MIF serum levels were detected by a commercially enzyme-linked immunosorbent assay (ELISA) (R&D SYSTEMS Human MIF DuoSet) kit according the protocol provided by the manufacturer with a lower limit of 31.3 pg/mL. Serum levels of MIF in sham-operated, HS and ISO-1 treated rats from the acute model of HS were quantified using a commercially available ELISA kit (Cusabio Biotech, Wuhan, China) with a lower limit of 62.5 pg/mL by following the manufacturer/product specific protocol. Detection occurred at 450 nm and 540 nm using iMark® microplate absorbance reader (BioRad).

### Western Blot Analysis

Semi-quantitative immunoblot analysis were carried out in liver and kidney tissue samples as previously described ^31^. The following antibodies were used: rabbit anti-NF-κB, rabbit anti-IKKα/β, rabbit anti-Ser176/180 IKKα/β, rabbit anti NLRP3 inflammasome (from Abcam) and mouse anti-caspase 1 (p20) (from Adipogen). Detailed description of the method can be found in the supplemental section.

### Statistical Analysis

All data in text and figures are expressed as box and whiskers plotted from min to max of n observations, where n represents the number of animals studied. Measurements obtained from the sham, control and ISO-1 treated groups were analyzed by one-way ANOVA followed by a Bonferroni’s *post-hoc* test on GraphPad Prism 8.0 (GraphPad Software, Inc., La Jolla, CA, USA). The distribution of the data was verified by Shapiro-Wilk normality test and the homogeneity of variances by Bartlett test. When necessary, values were transformed into logarithmic values to achieve normality and homogeneity of variances. To investigate the relationship between the variables Pearson correlation r was performed. Differences were considered to be statistically significant when p<0.05.

## Results

### MIF levels in serum of polytrauma patients are elevated and associated with longer stay on ICU and in hospital

Patients with blunt trauma have increased levels of cytokines including MIF and this correlates with poor outcome 15. To investigate the role of MIF in trauma, a total of 208 patients with a median age of 47.0 (31-60) were included in the study. Detailed patients characteristics can be found in Supplemental Table 2. MIF levels were measured in serum at different time points. Polytrauma patients showed significant increased MIF levels (12398 ± 1262 pg/ml) on arrival in the emergency room compared to 2866.9 ± 377.8 pg/mL after 2 days, to 2335.70 ± 203.4 pg/mL after 5 days and to 2114.6 ± 165.3 pg/mL after 7 days (all p<0.001) (Figure 1A). Furthermore, we found a weak positive correlation between MIF levels on arrival in the emergency room and both ICU (r=0.26, n=198, p<0.01) and hospital days (r=0.22, n=199, p<0.01). MIF levels on arrival in the emergency room were not correlated with baseline characteristics such as age (r=-0.05, n=200, p=0.49) and sex (r=-0.07, n=200, p=0.3) and only weakly correlated with the ISS score (r=0.15, n=177, p<0.05) (Figure 1B).

**Figure 1:**
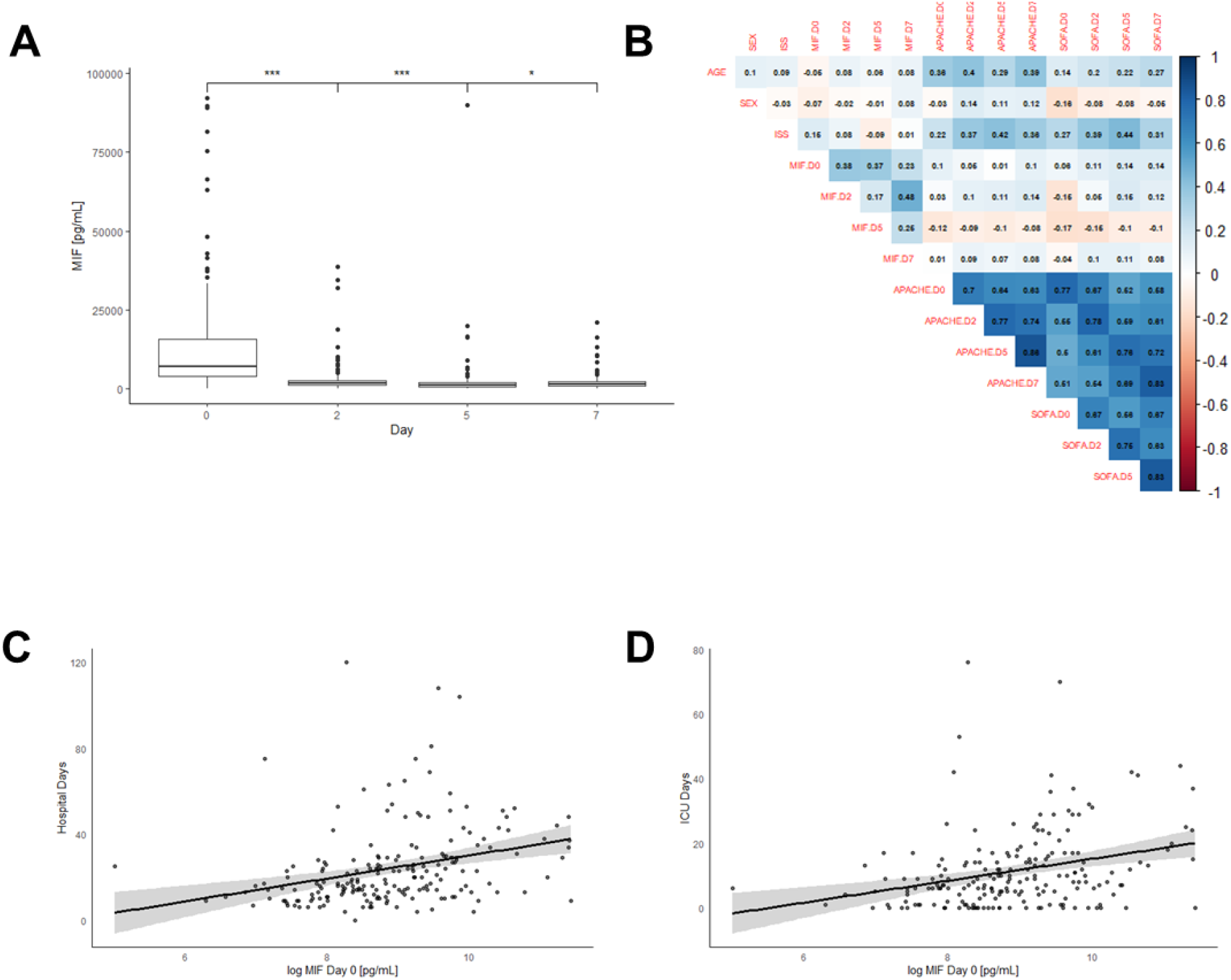
MIF serum levels are elevated in polytrauma patients. **(A)** Boxplot of MIF serum levels in trauma patients (n = 208) at different time points: Day 0 = Emergency room, Day 2 = after 48 hours; Day 5 = 120 hours; Day 7 = after 168 hours. Data shown as scattered dot plot. **(B)** Heatmap for correlations between MIF levels and baseline characteristics sex, age, SOFA, ISS and APACHE score. Scatter plot for **(C)** hospital stay and **(D)** ICU stay and MIF levels at arrival time in emergency room.

### MIF levels are elevated in serum of rats after induction of acute HS

We measured elevated serum levels of MIF in patients with trauma-hemorrhage, next we investigated to explore whether hemorrhage alone (in the absence of physical trauma) is sufficient to drive increases in MIF. To address this question, we used a model of severe hemorrhage followed by resuscitation in the rat. When compared to sham-operated rats, hemorrhage followed by resuscitation resulted in a significant increase in MIF levels (p<0.05; Figure 2A). Although ISO-1 has been reported to inhibit the effects, rather than the formation, of MIF *in vivo*, we report here that treatment of HS-rats with ISO-1 resulted in a significantly lower MIF level when compared to HS-rats treated with vehicle (p<0.05; Figure 2A). Administration of ISO-1 to sham-operated rats had no effect on MIF levels (p>0.05; Figure 2A).

**Figure 2:**
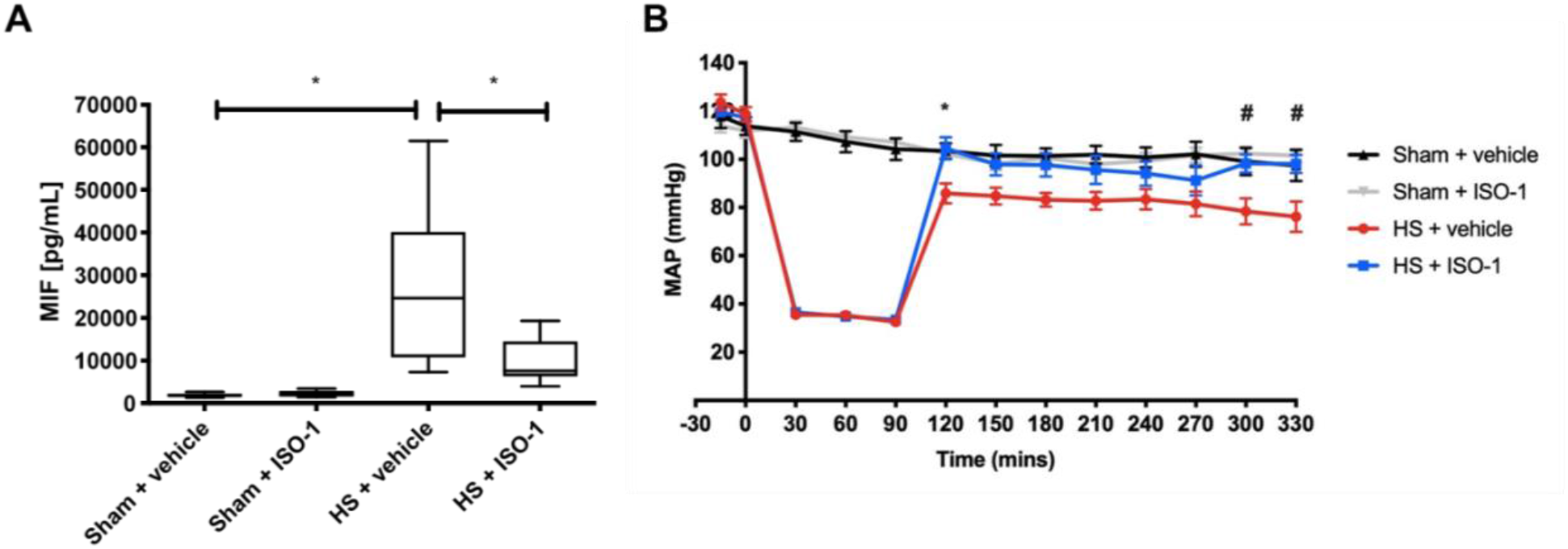
Treatment with ISO-1reduces MIF serum levels and improves HS-induced circulatory failure in an acute HS model. **(A)** MIF levels were detected by ELISA in sham-operated treated with vehicle or ISO-1 and HS rats treated with vehicle or ISO-1. Serum levels of MIF were significantly increased in the HS rats compared to sham. Treatment with ISO-1 had no significant effect on the rise in MIF-levels caused by HS. Data are expressed as box and whiskers plotted from min to max of ten animals per group. **(B)** Mean arterial pressure (MAP) was measured from the completion of surgery to the termination of the experiment for all groups. ISO-1 treatment lessened the decrease in MAP post-resuscitation compared to HS-rats treated with vehicle. Statistical analysis was performed using one-way ANOVA followed by a Bonferroni’s *post-hoc* test. ^*^*p* < 0.05 sham + vehicle vs. HS + vehicle; ^#^*p* < 0.05 HS + vehicle vs. HS + ISO-1.

### Treatment with ISO-1 improves HS-induced circulatory failure in an acute HS model

Hemorrhage reduces the circulating blood volume and cardiac output. To investigate the effects of MIF-inhibition with ISO-1 on circulatory failure, mean arterial blood pressure (MAP) was measured from the completion of surgery to the termination of the experiment. Baseline MAP values were similar amongst all four groups. Rats subjected to HS demonstrated a decline in MAP which was ameliorated by resuscitation, but MAP still remained lower than that of sham-operated rats during resuscitation (at the equivalent time points, Figure 2B). When compared to sham-operated rats, HS-rats treated with vehicle exhibited a more pronounced decrease in MAP over time post-resuscitation (p<0.05; Figure 2B). In contrast, MAP of HS-rats treated with ISO-1 was significantly higher than those in HS-rats treated with vehicle 4 h post-resuscitation (p<0.05; Figure 2B). Administration of ISO-1 to sham-operated rats had no significant effect on MAP (p>0.05; Figure 2B).

### Treatment with ISO-1 attenuates HS-induced organ damage in an acute HS model

We demonstrated that treatment with ISO-1 improves HS-induced circulatory failure in an acute HS model, we next explored whether pharmacological intervention with the MIF tautomerase inhibitor ISO-1 attenuates the MODS associated with HS in rats. Rats subjected to HS displayed a significant change in parameters evaluating organ injury and dysfunction when compared to sham-operated rats (Figure 3). The increases in serum urea (p<0.05; Figure 3A) and creatinine (p<0.05; Figure 3B) and the decrease in creatinine clearance (p<0.05; Figure 3C) indicated the development of renal dysfunction. The significant increases in both ALT (p<0.05; Figure 3D) and AST (p<0.05; Figure 3E) indicated the development of hepatic injury while the increases in amylase (p<0.05; Figure 3F) and CK (p<0.05; Figure 3G) denoted pancreatic and neuromuscular injury respectively. The significant increase in LDH (p<0.05; Figure 3H) confirmed tissue injury had occurred, but did not identify in which specific organ. The increase in lactate (p<0.05; Figure 3I) indicated decreased transport of oxygen to the tissues developing from the state of hypoperfusion. Treatment with ISO-1 resulted in a significant decrease of creatinine (p<0.05; Figure 3B and C), hepatic, pancreatic, neuromuscular and general tissue injury as shown by the decrease in serum parameter values creatinine (all p<0.05; Figure 3C-I). Administration of ISO-1 to sham-operated rats had no significant effect on any parameters measured (p>0.05; Figure 3).

**Figure 3:**
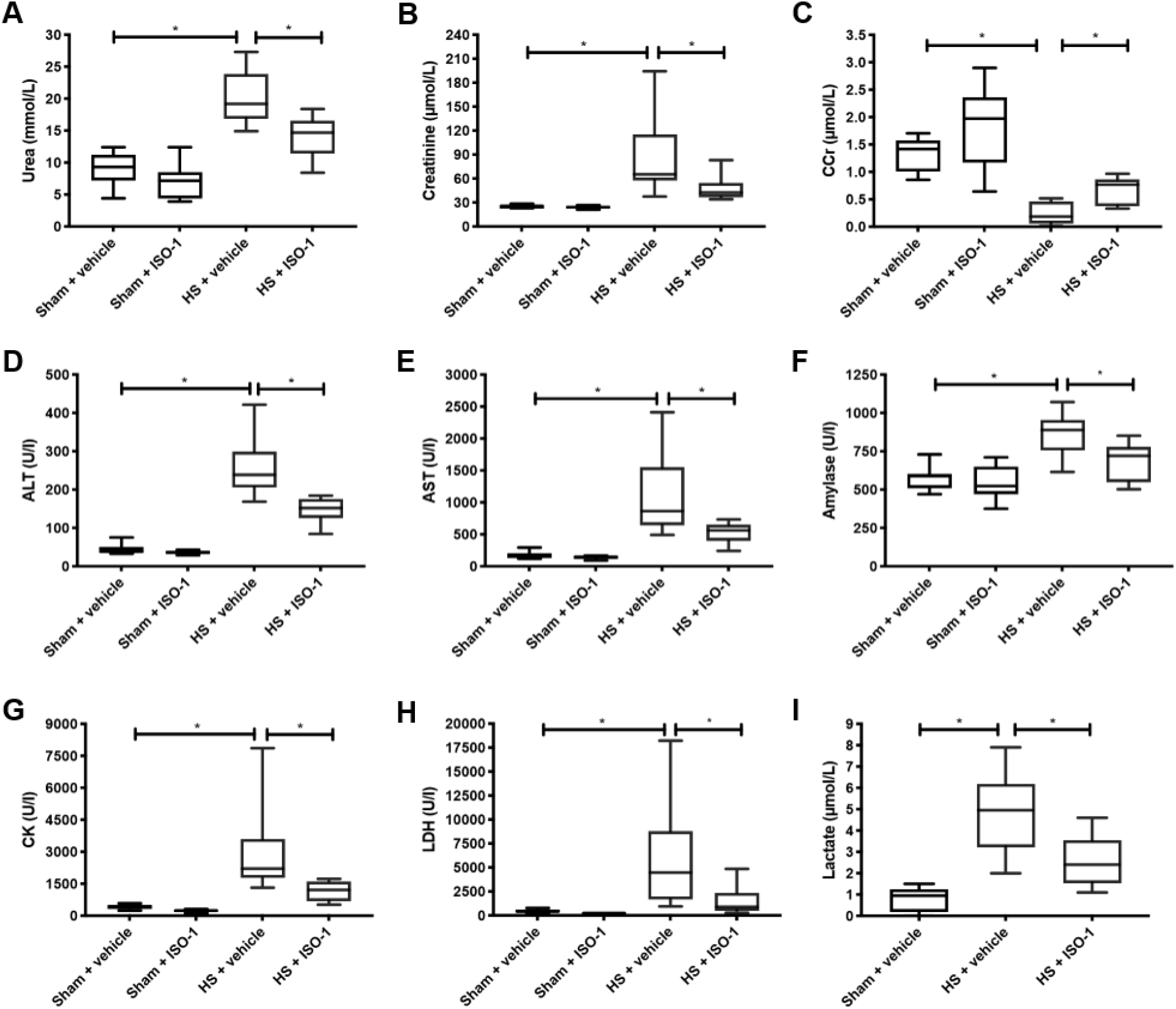
Treatment with ISO-1 attenuates HS-induced organ damage in an acute HS model. Rats were subjected to hemorrhagic shock (HS) and 4 h after resuscitation, levels of serum (**A**) urea, (**B**) creatinine and (**C**) creatinine clearance (CCr) as a measure of renal function; (**D**) alanine aminotransferase (ALT) and (**E**) aspartate aminotransferase (AST) as a measure of liver injury; (**F**) amylase as a measure of pancreatic injury; (**G**) creatine kinase (CK) as a measure of neuromuscular injury; (**H**) lactate dehydrogenase (LDH) as a non-specific measure of tissue injury; and (**I**) lactate as a measure of tissue hypoxia were determined. Sham-operated rats were used as control. All parameters were increased (with the exception of CCr which decreased) following HS compared to sham. ISO-1 treatment improved the parameter levels. Data are expressed as box and whiskers plotted from min to max of 10 animals per group. Statistical analysis was performed using one-way ANOVA followed by a Bonferroni’s post-hoc test. ^*^*p* < 0.05 denoted statistical significance.

### Treatment with ISO-1 attenuates hepatic and renal NF-κB activation in an acute HS model

The effect of MIF inhibition on the activation of key inflammatory signaling cascades, including pathways leading to NF-κB activation, were investigated in the liver and kidney. When compared to sham-operated rats, HS-rats treated with vehicle had significant increases in the phosphorylation of IKKα/β at Ser^176/180^ (p<0.05; Figures 4A and 4B) and the translocation of p65 to the nucleus (p<0.05; Figures 4C and 4D). Treatment with ISO-1 in HS-rats significantly attenuated the increases in hepatic and renal phosphorylation of IKKα/β at Ser^176/180^ (p<0.05; Figures 4A and 4B) and the nuclear translocation of p65 (p<0.05; Figures 4C and 4D).

**Figure 4:**
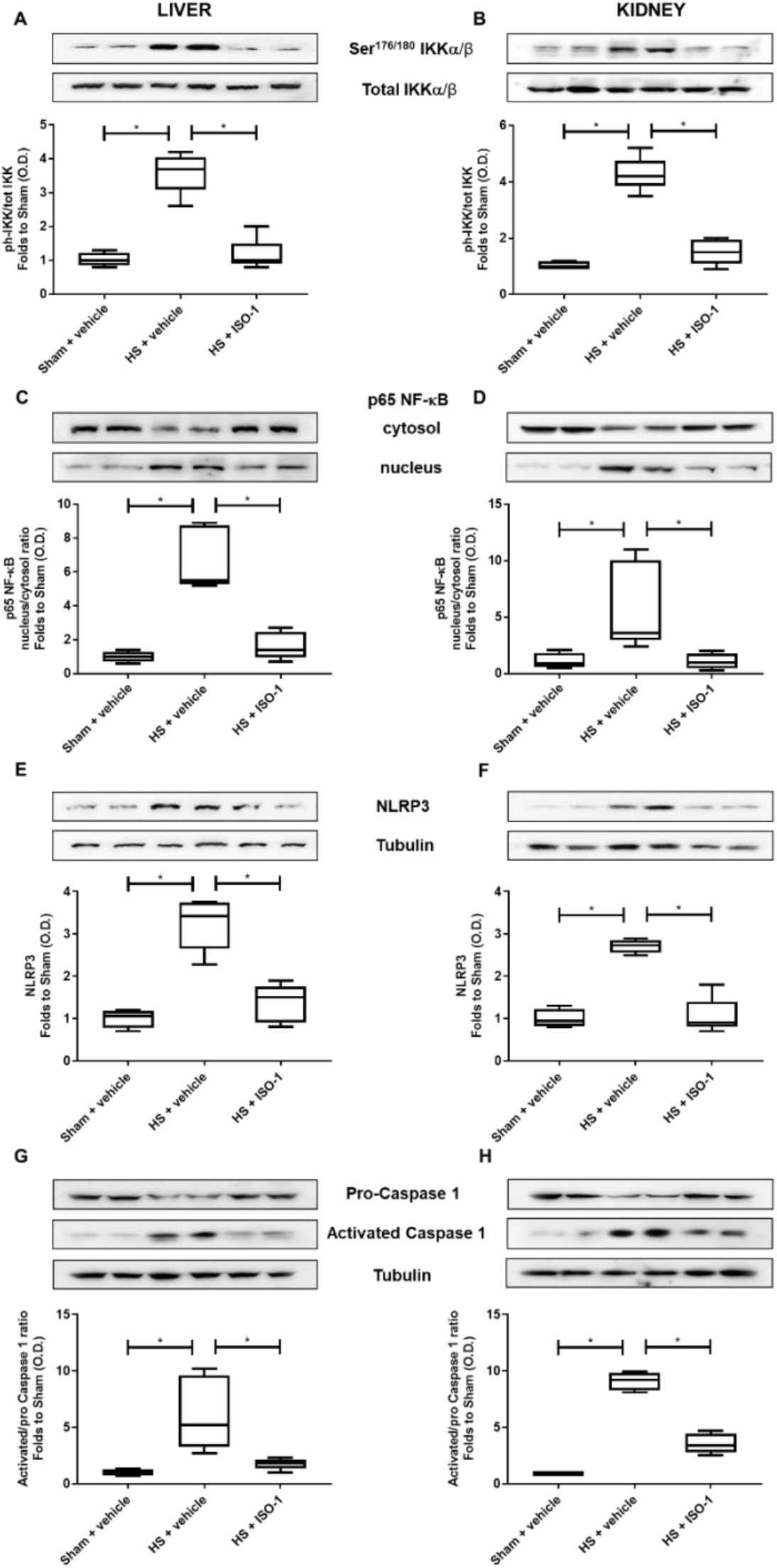
Treatment with ISO-1 attenuates NF-κB and NLRP3 activation in an acute HS model. **(A, B)** The phosphorylation of IKKα/β at Ser^176/180^, **(C, D)** the nuclear translocation of p65, **(E, F)** the activation of NLRP3 and **(G, H)** the cleavage of pro-caspase 1 of vehicle and ISO-1 treated rats were determined by Western blotting in the liver and kidney. Protein expression was measured as relative optical density (O.D.) and normalized to the sham band. Data are expressed as box and whiskers plotted from min to max of four to five animals per group. Statistical analysis was performed using one-way ANOVA followed by a Bonferroni’s post-hoc test. *p < 0.05 denoted statistical significance.

### Treatment with ISO-1 attenuates hepatic and renal NLRP3 inflammasome activation in an acute HS model

Having discovered that ISO-1 significantly reduced the activation of NF-κB in the liver and kidney of rats subjected to HS, we next analyzed the potential involvement of expression and activation of the NLRP3 inflammasome complex. When compared to sham-operated rats, HS-rats treated with vehicle exhibited a significantly increased expression of the NLRP3 inflammasome (p<0.05; Figures 4E and F) and cleavage of pro-caspase 1 to caspase 1 (p<0.05; Figures 4G and H). Treatment with ISO-1 in HS-rats significantly inhibited the hepatic and renal expression of NLRP3 (p<0.05; Figures 4E and F) and cleavage of pro-caspase 1 to caspase 1 (p<0.05; Figures 4G and H), suggestive of ISO-1protective effects against HS-induced overexpression and activation of the NLRP3 inflammasome complex

### Treatment with ISO-1 improves HS-induced circulatory failure in a chronic HS model

Having demonstrated that treatment with ISO-1 improved blood pressure in an acute HS rat model, we wished to determine whether ISO-1 would still be effective in a HS model in which the resuscitation period is prolonged to 24 h. When compared to sham-operated rats, HS-rats treated with vehicle had significantly lower values of MAP at 24 h post-resuscitation (p<0.05; Figure 5A); highlighting that cardiovascular dysfunction was still present. In contrast, the MAP values of HS-rats treated with ISO-1 were significantly higher at the 24 h time point than those of vehicle treated rats (p<0.05; Figure 5A). There were no significant differences in HR between any of the three groups investigated (p>0.05; Figure 5B).

**Figure 5:**
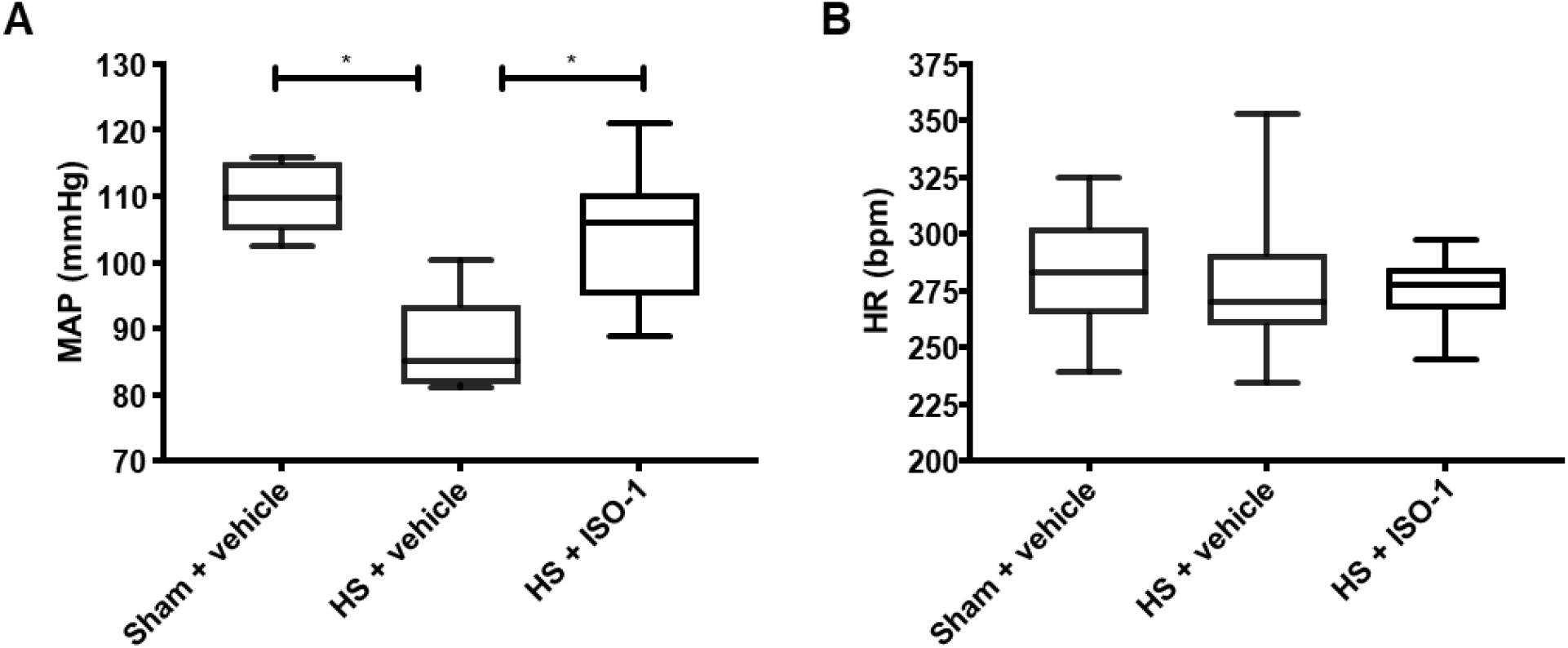
Treatment with ISO-1 improves HS-induced cardiac dysfunction in a chronic HS model. (**A**) HS resulted in a lower MAP compared to sham-operated rats, but this was improved with ISO-1 treatment. (**B**) There was no significant change in HR. Data are expressed as box and whiskers plotted from min to max. Sham + vehicle (n = 6), HS + vehicle (n = 12) and HS + ISO-1 (n = 12). Statistical analysis was performed using one-way ANOVA followed by a Bonferroni’s post-hoc test. *p < 0.05 denoted statistical significance.

### Treatment with ISO-1 attenuates HS-induced organ damage in a chronic HS model

Having shown that treatment with ISO-1 ameliorated the MODS associated with HS in an acute HS rat model, we examined whether efficacy was present 24 h post-resuscitation in a chronic HS model. As with the acute HS model, rats subjected to chronic HS displayed a significant change in parameters evaluating organ injury and dysfunction, when compared to sham-operated rats (Figure 6). The increases in serum urea (p<0.05; Figure 6A) and creatinine (p<0.05; Figure 6B) indicated the development of renal dysfunction. The increases in both ALT (p<0.05; Figure 6C) and AST (p<0.05; Figure 6D) indicated the development of hepatic injury while the increase in lipase (p<0.05; Figure 6E) denoted pancreatic injury. The increase in LDH (p<0.05; Figure 6F) confirmed general tissue injury. Similarly, ISO-1 treatment significantly reduced the extent of renal, hepatic and tissue injury as shown by the decrease in serum parameter values (all p<0.05; Figure 6A-D and F). Treatment with ISO-1 did not result in a decrease in lipase (Figure 6E).

**Figure 6:**
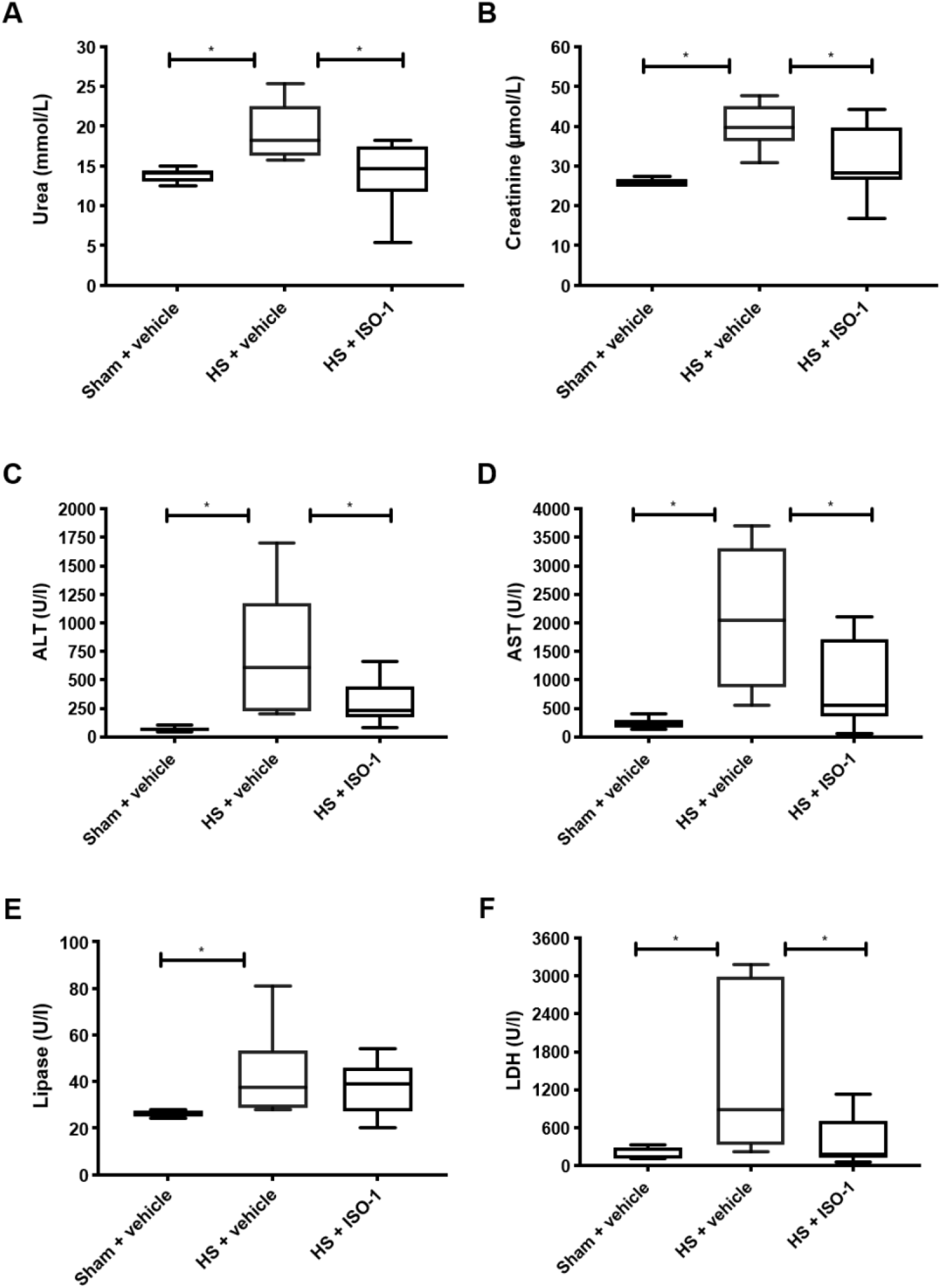
Treatment with ISO-1 attenuates HS-induced organ damage in a chronic HS model. Rats were subjected to hemorrhagic shock (HS) and 24 h after resuscitation, levels of serum (**A**) urea and (**B**) creatinine as a measure of renal function; (**C)** alanine aminotransferase, (ALT) and (**D**) aspartate aminotransferase (AST) as a measure of liver injury; (**E**) lipase and (**F**) LDH as a measure of pancreatic injury. Sham-operated rats were used as control. All parameters were increased following HS compared to sham. ISO-1 treatment improved the parameter levels. Data are expressed as box and whiskers plotted from min to max. Sham + vehicle (n = 6), HS + vehicle (n = 12) and HS + ISO-1 (n = 12). Statistical analysis was performed using one-way ANOVA followed by a Bonferroni’s post-hoc test. *p < 0.05 denoted statistical significance.

### Treatment with ISO-1 attenuates Cluster of Differentiation (CD) 68+ cell infiltration in an acute HS model and myeloperoxidase activity in a chronic HS model

As HS causes macrophage infiltration into the lungs ^30^, we measured CD68+ positive cells as a marker for macrophage invasion using immunohistology. In HS-rats treated with vehicle, we found a significant increase of macrophages count (47.33 ± 8.75 per field) compared to sham-operated animals (21.75 ± 2.29 per field, p<0.05), while the macrophage counts in HS-rats treated with ISO-1 was lower (35.90 ± 4.30 per field; Supplemental Figure 2).

Having demonstrated that treatment with ISO-1 reduced the cell infiltration in the lung in an acute HS rat model, we next determined myeloperoxidase (MPO) activity in the lung and liver as an indicator of neutrophil infiltration. When compared to sham-operated rats, HS-rats treated with vehicle showed a significant increase in MPO activity in the lung (p<0.05; Figure 7A) and liver (p<0.05; Figure 7B). Treatment with ISO-1 in HS-rats significantly attenuated these rises in MPO activity (p<0.05; Figure 7).

**Figure 7:**
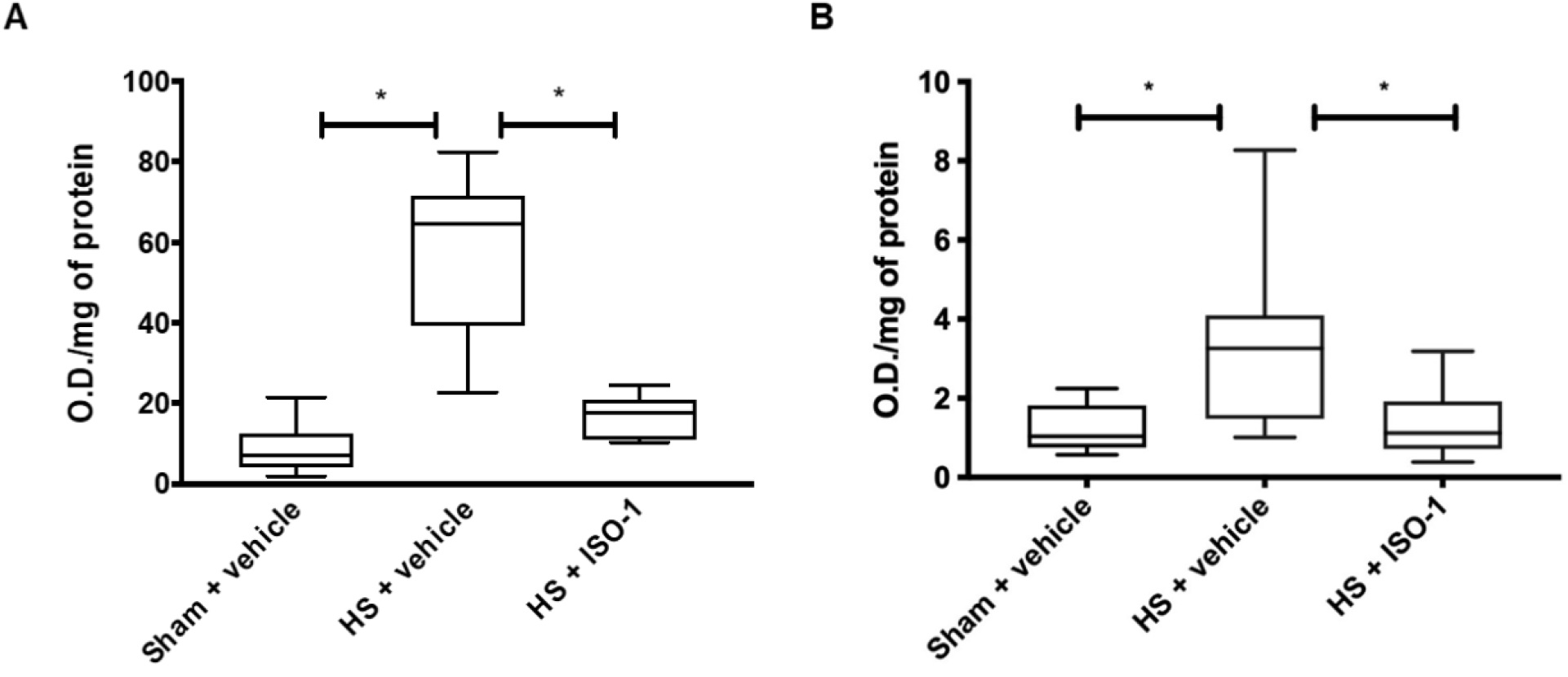
Treatment with ISO-1 attenuates pulmonary and hepatic myeloperoxidase activity in a chronic HS model. Myeloperoxidase activity in (**A**) lung and (**B**) liver were determined for vehicle and ISO-1 treated rats. Data are expressed as box and whiskers plotted from min to max. Sham + vehicle (n=6); HS + vehicle (n=12); HS + ISO-1 (n=12). Statistical analysis was performed using one-way ANOVA followed by a Bonferroni’s post-hoc test. *p<0.05 was considered statistically significant.

## Discussion

This study demonstrates for the first time a role of MIF in the pathophysiology of the organ injury and dysfunction caused by trauma/hemorrhage and indicates that inhibitors of MIF tautomerase activity may have potential in the therapy of the MODS after trauma and/or hemorrhage.

Although in polytrauma patients peak MIF levels were observed on admission and levels declined thereafter, they were still significantly elevated at day 7 after trauma (Figure 1A). MIF is a pleiotropic inflammatory cytokine with chemokine-like functions, that is rapidly released from pre-formed pools in the cytosol of several different cell types in response to various noxious stimuli such as infection, inflammation or hypoxia. MIF acts as a regulator of the initial immune response which may initiate and exacerbate various acute and chronic inflammatory disorders including acute respiratory distress syndrome and sepsis ^32, 33^.

Cho and colleagues showed that trauma patients with elevated MIF levels have longer ICU-stays than patients with lower or normal MIF levels ^34^. Indeed, we detected a significant correlation between elevated MIF levels in patients with trauma and the length of hospital stay (p<0.01) as well as the length of stay on ICU (p<0.01) (Figures 1C and 1D). These findings indicate that high MIF serum levels at time of admission are strongly predictive for a longer stay in ICU and hospital. However, we found only weak associations between MIF and ISS scores (Figure 1B). Thus, further studies are necessary to gain a better insight into the role of MIF in the pathophysiology of trauma and its complications including multi organ dysfunction syndrome.

To gain a better insight into the role of MIF in the pathogenesis of inflammation and organ injury/dysfunction associated with severe hemorrhage, we investigated the role of MIF in rat models of HS by inhibiting the tautomerase activity site of MIF with ISO-1. Indeed, there is good evidence that inhibition of the pro-inflammatory activity of MIF is highly beneficial in the treatment of many diseases, including sepsis ^33, 35^. Furthermore, Rosengren and colleagues discovered a strong correlation between the specific inhibition of MIF tautomerase activity by ISO-1 and suppression of the pro-inflammatory effects of MIF pro-inflammatory activities ^36^. A further study also showed, that the specific inhibition of MIF tautomerase activity in macrophages treated with LPS resulted in a decreased NF-κB activation and TNF production. Additionally, they demonstrated that ISO-1 treatment 24 h after induction of polymicrobial sepsis by cecal ligation and puncture was associated with a higher survival rate ^37^. Disruption of the active site by insertion of an alanine between Pro-1 and Met-2 abolishes the tautomerase activity of MIF and the resultant mutant is defective in the *in vitro* glucocorticoid counter-regulatory activity of MIF ^38^. Several other studies have supported the link between MIF bioactivity and this active site, whereas the exact biological function of this tautomerase remains speculative ^39^.

We report here that severe hemorrhage (for 90 min) followed by resuscitation in the rat caused a significant increase in the serum levels of MIF, the magnitude of which was similar to the one seen in trauma patients. This finding implies that hemorrhage (rather than trauma) is the main driver for the observed increase in MIF in rats and possibly also in humans. Furthermore, ISO-1 significantly attenuated the fall in blood pressure in the acute as well as the chronic HS (resuscitation for 24 h) model (Figure 2B and 5A). Thus, ISO-1 reduces the delayed vascular decompensation associated with HS ^40^. Rats subjected to acute HS had renal dysfunction, liver injury, pancreatic injury and lung inflammation (Figure 3 and 6). This organ injury/dysfunction and lung inflammation was attenuated in HS-rats (acute model) treated with ISO-1 (Figure 3 and 6). Even when the resuscitation period was increased to 24 h, animals subjected to HS (chronic model) still had a significant degree of organ dysfunction. This late MODS was also attenuated by treatment of rats with ISO-1 during the early resuscitation period (Figure 6A and D).

The lung inflammation caused by HS is associated with a significant recruitment of leukocytes to the lung. We analyzed the degree of macrophage infiltration (number of CD68-positive cells) in the acute HS model in the lung and the degree of neutrophil infiltration (MPO activity) in the chronic HS model in the lung and liver. We found a significant increase in CD68-positive cells after HS (acute model) and a significant increase in MPO activity in lung and liver (chronic HS-model). Treatment of HS-rats with ISO-1 did not significantly affect macrophage recruitment into the lung (Supplemental Figure 2). In contrast, treatment of HS-rats with ISO-1 attenuated the rise in MPO activity and, hence, the neutrophil recruitment in lung and liver (chronic HS-model) (Figure 7). Taken together, these findings imply that ISO-1 attenuates neutrophil, but not macrophage recruitment in HS. These findings can be explained by the chemokine like function of MIF, which orchestrates the activation and recruitment of leukocytes during immune surveillance and inflammation ^41^. MIF-mediated recruitment of mononuclear cells has been implicated in many diseases, however, its promigratory functions have been ascribed to its pleiotropic properties in cell activation.

Trauma results in increased translocation of NF-κB to the nucleus (in many organs and cells), which, in turn, drives the increased production of pro-inflammatory cytokines ^30^. Indeed, we found a significant increase in the degree of phosphorylation of IKKα/β at Ser176/180 and nuclear translocation of NF-κB subunit p65 in the liver and kidney in rats with acute HS (Figure 4A-D). Furthermore, we detected a significant decrease of phosphorylation of IKKα/β on Ser176/180 and nuclear translocation of NF-kB subunit p65 after treatment with ISO-1 in liver and kidney in rats with acute HS (Figure 4A-D). The present data are in line with previous findings indicating that MIF upregulates TLR4 expression resulting increased translocation of NF-κB into the nucleus ^42^. A previous study also indicates that ISO-1 inhibits TNF-release from macrophages isolated from LPS-treated mice^43^. HS and resuscitation (acute HS-model) also resulted in a significant increase in NLRP3 expression and activation resulting a significant increase caspase 1 formation in liver and kidney (Figure 4E-H). In contrast, treatment of HS-rats with ISO-1 resulted in a significant decrease of NLRP3 activation (Figure 4E-H). These results suggested that MIF (either directly or indirectly) drives the activation of the NLRP3 inflammasome that occurs after severe hemorrhage and resuscitation. These results are in line with previous studies which shows that MIF has a role in the activation of the NLRP3 inflammasome ^44, 45^.

## Conclusion

In conclusion, we demonstrate here for the first time that MIF is elevated on admission in patients with poly-trauma. MIF levels remain elevated for up to seven days after trauma and higher MIF serum levels (on arrival in the emergency room) are associated with a longer stays in the ICU and, indeed, the hospital. The finding that hemorrhage alone in rats resulted in a similar rise in MIF-levels than trauma/hemorrhage in patients supports the view that hemorrhage (organ ischemia) is the main driver for the elevations in MIF in animals and man with trauma/hemorrhage. Furthermore, we detected that the inhibition of the MIF tautomerase activity side with ISO-1 reduces the organ injury/dysfunction caused by severe hemorrhage and resuscitation in both an acute and a more chronic model of HS. Thus, ISO-1 (or other strategies that reduce the effects of MIF-1) may be a new therapeutic strategy for patients with trauma and severe hemorrhage. As trauma is a common cause of death and responsible for longer stays on hospital or ICU, it is very important to find a biomarker that can be measured on admission in the emergency room to identify patients who will need more care and resource, because of an expected prolonged stay in ICU and/or hospital. The results of this study suggested that MIF may have the potential to be such a biomarker able to identify patients who will have prolonged ICU and hospital stays after trauma and hemorrhage.

## Data Availability

The data that support the findings of this study are available from the corresponding author upon reasonable request.

## Abbreviations

ALT: alanine aminotransferase
AST: aspartate aminotransferase
CCr: creatinine clearance
CD: Cluster of Differentiation
CK: creatine kinase
DAMP: damage-associated molecular patterns
HR: heart rate
HS: hemorrhagic shock
ICU: intensive care unit
ISS: Injury Severity Score
LDH: lactate dehydrogenase
MAP: mean arterial pressure
MIF: macrophage migratory inhibition factor
MODS: multi organ dysfunction syndrome
MPO: myeloperoxidase

## Declarations

### Ethics approval and consent to participate

Blood samples of 208 patients were collected after written informed consent was obtained from either patient or a nominated legally authorized representative as approved by the ethical committee. Samples of the included subjects were collected between years 2010-2014 from University Hospital Frankfurt of Goethe-University and approved by an institutional ethics committee (number 312/10) in accordance with the declaration of Helsinki and following STROBE-guidelines ^26^.

For the acute HS model, all animal procedures were approved by the Animal Welfare Ethics Review Board (AWERB) of Queen Mary University of London and by the Home Office (License number PC5F29685).

For the chronic HS model, all animal care and experimental procedures were approved by the Committee for Animal Use in Research (License number 7396250219) of the Universidade Federal de Santa Catarina Institutional and are in accordance with the Brazilian Government Guidelines for Animal Use in Research (CONCEA).

All *in vivo* experiments are reported in accordance to ARRIVE guidelines.

### Consent for publication

Informed consent was obtained from all subjects involved in the study or their legal representatives.

### Conflict of interest

The authors declare no conflict of interest. The funders had no role in the design of the study; in the collection, analyses, or interpretation of data; in the writing of the manuscript, or in the decision to publish the results.

### Funding

NMP was funded by the William Harvey Research Foundation, HPR and FRMBO were funded by National Council for Scientific and Technological Development (CNPq) fellowship. This study was supported by the German Research Foundation to LM (DFG, MA 7082/3-1), to CS (DFG, STO 1099/8-1) and by an intramural grant to EZ (START 131/19), National Council for Scientific and Technological Development to RS (CNPq, Brazil, Grant 409018/2018-0).

## Author Contributions

Conception and design: NMP, LM, and CT; Animal experiments: NY, NMP, FRMBO, HPR and RS; Human sample analysis: LM, EZ, GM, IM, BR, DH, LS and CS; Animal sample analyses: NY, NMP, LM, CT, LS, EZ, FRMBO, HPR, SK, DC, MC, GFA and RS; Clinical study and patient data analyses: LM, EZ, CT, IM, BR, DH, LS, NMP and CS; Statistical analyses: NY, NMP, DH, LM and CT; Drafting the manuscript for important intellectual content: LS, LM, EZ, NMP and CT; All authors reviewed and approved the manuscript.

## Appendices

Supplemental Data.pdf

## Notes

### Competing Interest Statement

The authors have declared no competing interest.

### Author Declarations

Ethics approval and consent to participate Blood samples of 208 patients were collected after written informed consent was obtained from either patient or a nominated legally authorized representative as approved by the ethical committee. Samples of the included subjects were collected between years 2010-2014 from University Hospital Frankfurt of Goethe-University and approved by an institutional ethics committee (number 312/10) in accordance with the declaration of Helsinki and following STROBE-guidelines. Ethical approval was given to Department of Trauma, Hand and Reconstructive Surgery, University Hospital Frankfurt, Johann Wolfgang Goethe University, Theodor Stern Kai 7, Frankfurt D-60590, Germany. For the acute HS model, the Animal Welfare Ethics Review Board (AWERB) of Queen Mary University of London approved all experiments in accordance with the Home Office Guidance on the Operation of Animals (Scientific Procedure Act, 1986) published by Her Majesty's Stationary Office, and the Guide for the Care and Use of Laboratory Animals of the National Research Council. Work was conducted under UK Home Office Project License Number PC5F29685. Ethical approval was given to Professor Christoph Thiemermann, Department of Translational Medicine & Therapeutics, William Harvey Research Institute, Barts and The London School of Medicine and Dentistry, Queen Mary University of London, London, EC1M 6BQ, United Kingdom For the chronic HS model, all animal care and experimental procedures were approved by Universidade Federal de Santa Catarina Institutional Committee for Animal Use in Research (License number 7396250219) and are in accordance to the Brazilian Government Guidelines for Animal Use in Research (CONCEA). Ethical approval was given to Department of Pharmacology, Universidade Federal de Santa Catarina, SC, 88040-900, Brazil. All in vivo experiments are reported in accordance to ARRIVE guidelines. Consent for publication Informed consent was obtained from all subjects involved in the study or their legal representatives.

## References

1. Haagsma JA, Graetz N, Bolliger I, et al. The global burden of injury: Incidence, mortality, disability-adjusted life years and time trends from the global burden of disease study 2013. Injury Prevention 2016; 22(1):3–18.

2. Violence I. Injuries Violence. 2014.

3. Teixeira PGR, Inaba K, Hadjizacharia P, et al. Preventable or Potentially Preventable Mortality at a Mature Trauma Center. Journal of Trauma and Acute Care Surgery 2007; 63(6).

4. Dutton RP, Stansbury LG, Leone S, et al. Trauma mortality in mature trauma systems: Are we doing betterã an analysis of trauma mortality patterns, 1997-2008. Journal of Trauma-Injury, Infection and Critical Care 2010; 69(3):620–626.

5. Geeraedts LMG, Kaasjager HAH, van Vugt AB, et al. Exsanguination in trauma: A review of diagnostics and treatment options. Injury 2009; 40(1):11–20.

6. Sauaia A, Moore EE, Johnson JL, et al. Temporal trends of postinjury multiple-organ failure. Journal of Trauma and Acute Care Surgery 2014; 76(3):582–593.

7. Dewar D, Moore FA, Moore EE, et al. Postinjury multiple organ failure. Injury 2009; 40(9):912–918.

8. Lord JM, Midwinter MJ, Chen YF, et al. The systemic immune response to trauma: An overview of pathophysiology and treatment. The Lancet 2014; 384(9952):1455–1465.

9. Nerlich M, Kerschbaum M, Ernstberger A. Polytrauma-Management – präklinisches Handling und Schockraumversorgung: Aktualisierung 2017. Notfall und Rettungsmedizin 2017; 20(7):596–601.

10. Ulvik A, Kvåle R, Wentzel-Larsen T, et al. Multiple organ failure after trauma affects even long-term survival and functional status. Critical Care 2007; 11(5):1–8.

11. Eppensteiner J, Davis RP, Barbas AS, et al. Immunothrombotic activity of damage-associated molecular patterns and extracellular vesicles in secondary organ failure induced by trauma and sterile insults. Frontiers in Immunology 2018; 9(FEB):1–14.

12. Relja B, Mörs K, Marzi I. Danger signals in trauma. European Journal of Trauma and Emergency Surgery 2018; 0(0):0–0.

13. Chen GY, Nuñez G. Sterile inflammation: Sensing and reacting to damage. Nature Reviews Immunology 2010; 10(12):826–837.

14. Halbgebauer R, Braun CK, Denk S, et al. Hemorrhagic shock drives glycocalyx, barrier and organ dysfunction early after polytrauma. Journal of Critical Care 2018; 44:229–237.

15. Jackman RP, Utter GH, Muench MO, et al. Distinct roles of trauma and transfusion in induction of immune modulation post-injury. Transfusion 2013; 52(12):2533–2550.

16. Manson J, Thiemermann C, Brohi K. Trauma alarmins as activators of damage-induced inflammation. British Journal of Surgery 2012; 99(SUPPL. 1):12–20.

17. Lue H, Dewor M, Leng L, et al. Activation of the JNK signalling pathway by macrophage migration inhibitory factor (MIF) and dependence on CXCR4 and CD74. Cell Signal 2011; 23(1):135–44.

18. Calandra T. Protection from septic shock by neutralizing of macrophage migration inhibitory factor. Nature Medicine 2000.

19. Emonts M, Sweep FCGJ, Grebenchtchikov N, et al. Association between High Levels of Blood Macrophage Migration Inhibitory Factor, Inappropriate Adrenal Response, and Early Death in Patients with Severe Sepsis. Clinical Infectious Diseases 2007; 44(10):1321–1328.

20. Hayakawa M, Katabami K, Wada T, et al. Imbalance between macrophage migration inhibitory factor and cortisol induces multiple organ dysfunction in patients with blunt trauma. Inflammation 2011; 34(3):193–197.

21. Gombert A, Stoppe C, Foldenauer AC, et al. Macrophage Migration Inhibitory Factor Predicts Outcome in Complex Aortic Surgery. Int J Mol Sci 2017; 18(11).

22. Al-Abed Y, VanPatten S. MIF as a disease target: ISO-1 as a proof-of-concept therapeutic. Future medicinal chemistry 2011; 3(1):45–63.

23. Al-Abed Y, Dabideen D, Aljabari B, et al. ISO-1 binding to the tautomerase active site of MIF inhibits its pro-inflammatory activity and increases survival in severe sepsis. Journal of Biological Chemistry 2005; 280(44):36541–36544.

24. Lubetsky JB, Dios A, Han J, et al. The tautomerase active site of macrophage migration inhibitory factor is a potential target for discovery of novel anti-inflammatory agents. Journal of Biological Chemistry 2002.

25. Yang D-B, Yu W-H, Dong X-Q, et al. Serum macrophage migration inhibitory factor concentrations correlate with prognosis of traumatic brain injury. Clinica chimica acta; international journal of clinical chemistry 2017; 469:99–104.

26. von Elm E, Altman DG, Egger M, et al. The Strengthening the Reporting of Observational Studies in Epidemiology (STROBE) statement: guidelines for reporting observational studies. Bull World Health Organ 2007; 85(11):867–72.

27. Mörs K, Braun O, Wagner N, et al. Influence of gender on systemic IL-6 levels, complication rates and outcome after major trauma. Immunobiology 2016.

28. Baker SP, O’Neill B, Haddon W, Jr., et al. The injury severity score: a method for describing patients with multiple injuries and evaluating emergency care. J Trauma 1974; 14(3):187–96.

29. Sordi R, Nandra KK, Chiazza F, et al. Artesunate Protects Against the Organ Injury and Dysfunction Induced by Severe Hemorrhage and Resuscitation. Ann Surg 2017; 265(2):408–417.

30. Yamada N, Martin LB, Zechendorf E, et al. Novel Synthetic, Host-defense Peptide Protects Against Organ Injury/Dysfunction in a Rat Model of Severe Hemorrhagic Shock. Ann Surg 2018; 268(2):348–356.

31. Collino M, Pini A, Mugelli N, et al. Beneficial effect of prolonged heme oxygenase 1 activation in a rat model of chronic heart failure. Dis Model Mech 2013; 6(4):1012–20.

32. Donnelly SC, Haslett C, Reid PT, et al. Regulatory role for macrophage migration inhibitory factor in acute respiratory distress syndrome. Nat Med 1997; 3(3):320–3.

33. Calandra T, Roger T. Macrophage migration inhibitory factor: a regulator of innate immunity. Nature Reviews Immunology 2003; 3(10):791–800.

34. Cho YD, Choi SH, Kim JY, et al. Macrophage migration inhibitory factor levels correlate with an infection in trauma patients. Ulus Travma Acil Cerrahi Derg 2017; 23(3):193–198.

35. Riedemann NC, Guo RF, Gao H, et al. Regulatory role of C5a on macrophage migration inhibitory factor release from neutrophils. J Immunol 2004; 173(2):1355–9.

36. Rosengren E, Bucala R, Aman P, et al. The immunoregulatory mediator macrophage migration inhibitory factor (MIF) catalyzes a tautomerization reaction. Mol Med 1996; 2(1):143–9.

37. Al-Abed Y, Dabideen D, Aljabari B, et al. ISO-1 Binding to the Tautomerase Active Site of MIF Inhibits Its Pro-inflammatory Activity and Increases Survival in Severe Sepsis*. Journal of Biological Chemistry 2005; 280(44):36541–36544.

38. Lubetsky JB, Dios A, Han J, et al. The tautomerase active site of macrophage migration inhibitory factor is a potential target for discovery of novel anti-inflammatory agents. J Biol Chem 2002; 277(28):24976–82.

39. Swope M, Sun HW, Blake PR, et al. Direct link between cytokine activity and a catalytic site for macrophage migration inhibitory factor. Embo j 1998; 17(13):3534–41.

40. Thiemermann C, Szabó C, Mitchell JA, et al. Vascular hyporeactivity to vasoconstrictor agents and hemodynamic decompensation in hemorrhagic shock is mediated by nitric oxide. Proceedings of the National Academy of Sciences 1993; 90(1):267–271.

41. Bernhagen J, Krohn R, Lue H, et al. MIF is a noncognate ligand of CXC chemokine receptors in inflammatory and atherogenic cell recruitment. Nat Med 2007; 13(5):587–96.

42. Roger T, David J, Glauser MP, et al. MIF regulates innate immune responses through modulation of Toll-like receptor 4. Nature 2001; 414(6866):920–4.

43. Al-Abed Y, Dabideen D, Aljabari B, et al. ISO-1 binding to the tautomerase active site of MIF inhibits its pro-inflammatory activity and increases survival in severe sepsis. J Biol Chem 2005; 280(44):36541–4.

44. Lang T, Lee JPW, Elgass K, et al. Macrophage migration inhibitory factor is required for NLRP3 inflammasome activation. Nature Communications 2018; 9(1):2223.

45. Shin MS, Kang Y, Wahl ER, et al. Macrophage Migration Inhibitory Factor Regulates U1 Small Nuclear RNP Immune Complex-Mediated Activation of the NLRP3 Inflammasome. Arthritis Rheumatol 2019; 71(1):109–120.

