## Supplemental Data for "The inhibition of Macrophage Migration Inhibitory Factor by ISO-1 attenuates trauma-induced multi organ dysfunction in rats"

Dr Lukas Martin, MD PhD MHBA

Uniklinik RWTH Aachen

Department of Intensive Care and Intermediate Care

Pauwelsstraße 30

52074 Aachen, Germany

**Running head: ISO-1 attenuates trauma-induced MOF**

**Supplemental Methods**

*Acute Hemorrhagic Shock Model*

Rats were anesthetized with sodium thiopentone (120 mg/kg i.p. initially and 10 mg/kg i.v. for maintenance as needed). Cannulation with polyethylene catheters (Smiths Medical International Ltd., Kent, UK) of the trachea for facilitation of spontaneous breathing (internal diameter [ID] 1.67 mm), left femoral artery for recording of the mean arterial pressure (MAP) (ID 0.40 mm), left carotid artery for blood withdrawal (ID 0.58 mm) and right jugular vein for fluid and drug administration (ID 0.40 mm) was performed. To prevent tissue desiccation, swabs moistened with saline were placed over the surgery incision sites. Body temperature was monitored by a rectal probe thermometer and maintained at 37°C ± 0.3°C by means of a homoeothermic blanket system (Harvard Apparatus). Upon the completion of surgery, the MAP was allowed to stabilize for 15 min. Blood was then withdrawn (up to 1 mL/min into heparinized syringes containing 100 IU/mL heparin mixed with normal saline) through the cannula inserted in the carotid artery in order to achieve a fall in MAP to 35 ± 5 mmHg, which was recorded with a pressure transducer (attached to the femoral artery cannula, 844-31 Memscap, Durham, USA) and coupled to a PowerLab 8/30 data acquisition system (AD Instruments Pty Ltd., Castle Hill, Australia). Thereafter, MAP was maintained at 35 ± 5 mmHg for a period of 90 min either by further withdrawal of blood during the compensation phase or administration of the shed blood during the decompensation phase. At 90 min after initiation of hemorrhage (or when 25% of the shed blood had to be re-injected to sustain MAP at 35 ± 5 mmHg), resuscitation through the jugular vein was performed with the remaining shed blood (mixed with 100 IU/mL heparinized saline) over a period of 5 min plus a volume of Ringer's lactate identical to the volume of shed blood. Treatment or vehicle was also administered intravenously. One hour after resuscitation, an infusion of Ringer's lactate (1.5 mL/kg/h) was started as fluid replacement and it was maintained throughout the experiment for a total of 3 h. During the final 3 h, urine was collected through a catheter placed in the bladder to estimate the creatinine clearance. Under deep anesthesia, the heart was removed to terminate the experiment 4 h after resuscitation. Sham-operated rats were used as control and underwent identical surgical procedures, but without hemorrhage or resuscitation.

Rats were treated with either ISO-1 (25 mg/kg) or its vehicle (5% DMSO + 95% Ringer's lactate). An initial bolus of ISO-1 treatment (10 mg/kg) or vehicle was administered intravenously immediately after resuscitation. 30 min after resuscitation, an infusion of ISO-1 (3.75 mg/kg/h, i.v.) or vehicle was started and maintained throughout the experiment for a total of 4 h.

*Sample Collection - Acute Hemorrhagic Shock Model*

Rats remained anesthetized with sodium thiopentone (120 mg/kg i.p.) before sacrifice. Up to 8 mL of blood was taken from the right ventricle of the heart via cardiac puncture into non-heparinized 5 mL syringes and immediately decanted into 1.1 mL serum gel tubes (Sarstedt, Germany). The blood was centrifuged (10000 g for 5 min) to obtain the serum, which was subsequently stored at -80 °C until analysis. Organs (heart, lungs, liver and kidneys) were excised of which one section was snap frozen in liquid nitrogen and stored at -80 °C, and another section was placed in 10 % formalin for 24-48 h; followed by transfer to 70 % ethanol until further analysis. All organ injury/dysfunction parameters (urea, creatinine, alanine aminotransferase [ALT], aspartate aminotransferase [AST], creatine kinase [CK], amylase and lactate dehydrogenase [LDH]) in the serum were measured in a blinded fashion by a clinical pathology diagnostic laboratory (MRC Harwell Institute, Oxfordshire, UK).

*Chronic Hemorrhagic Shock Model*

Thirty rats were randomized into three groups: Sham + vehicle (n = 6); HS + vehicle (n = 12); HS + ISO-1 (25 mg/kg; n = 12). At 15 min prior to anesthesia, analgesia with tramadol (10 mg/kg i.p.) was administered. Rats were then anesthetized with ketamine-xylazine (ketamine,

100 mg/kg; xylazine, 10 mg/kg i.m. initially and 100 µL ketamine i.p. for maintenance as needed). Cannulation with polyethylene catheters of the left femoral artery and left femoral vein was performed. Body temperature was monitored by a digital ear thermometer and maintained at 36.5 °C ± 0.5 °C by means of a homoeothermic blanket system (Harvard Apparatus). Upon completion of surgery, the MAP was allowed to stabilize for 15 min. Blood was then withdrawn (up to 1 mL/min into heparinized syringes containing 100 IU/mL heparin mixed with normal saline) through the cannula inserted in the femoral artery in order to achieve a fall in MAP to 40 ± 2 mmHg, which was recorded with a pressure transducer (attached to the femoral artery cannula) and coupled to a PowerLab 8/30 (AD Instruments Pty Ltd., Castle Hill, Australia). Thereafter, MAP was maintained at 40 ± 2 mmHg for a period of 90 min either by further withdrawal of blood or administration of the shed blood. At 90 min after initiation of hemorrhage (or when 25 % of the shed blood had to be re-injected to sustain MAP at 40 ± 2 mmHg), resuscitation through the femoral vein was performed with the remaining shed blood over a period of 5 min plus 1.5 mL/kg Ringer's lactate (i.v.). An initial bolus of ISO-1 treatment (12.5 mg/kg) or vehicle was then administered intraperitoneally. Sham-operated rats were used as control and underwent identical surgical procedures, but without hemorrhage or resuscitation. At 20 min after resuscitation was completed, the catheters were removed, the femoral vessels were ligated and the incision was closed with sutures. Rats were allowed to recover from the anesthesia and 12 h later given tramadol (5 mg/kg i.p.) and a second dose of ISO-1 (12.5 mg/kg) or vehicle.

*Quantification of myeloperoxidase activity*

Lung and liver tissue samples from the chronic HS model were pulverized in liquid nitrogen with a pestle and mortar and homogenized in 80 mM phosphate-buffered saline (PBS), pH 5.4, containing 0.5% hexadecyltrimethylammonium bromide. The homogenate was then centrifuged at 13,000 x g at 4°C for 10 min and the supernatant was assayed for myeloperoxidase (MPO) activity by measuring the $H_2O_2$-dependent oxidation of 3,3,5,5-tetramethylbenzidine (TMB). MPO activity was determined colorimetrically using an ultra-microplate reader (EL 808, BioTek Instruments, INC, USA) set to measure absorbance at 650 nm. Total protein content in the homogenate was estimated using the BCA assay (Thermo Fisher Scientific, Rockford, IL), according to the manufacturer's instructions. MPO activity was expressed as optical density at 650 nm per mg of protein.

*Immunostaining*

Lung tissue was extracted at the end of the experiment and kept in formalin for 24 h to 48 h at room temperature, before it was transferred to 70% ethanol. Afterwards the tissue was embedded in paraffin and sectioned. The slides were then deparaffinized and rehydrated with xylene and ascending concentrations of ethanol.  The next step included an antigen retrieval to unmask unspecific antigen bindings. Afterwards followed incubation with rabbit anti-CD68 antibody ED1 (1:2000 in TBS with 1% BSA + 10% rabbit serum; catalog no. MCA341R; AbD Serotec) for 16 h at 4°C. The sections were then incubated with labelled polymer-HRP antibody (ab236469 - Rabbit specific HRP/DAB Detection IHC Detection Kit - Micro-polymer) for 15 minutes. Afterwards the slides were counterstained with Harris hematoxylin solution, dehydrated and mounted on slides. Images were developed using a NanoZoomer Digital Pathology Scanner (Hamamatsu Photonics K.K. Japan). NDP Viewer software was used for analyzation. CD68 positive cells were counted in 10 randomly selected fields (300 μm) in a double-blinded manner by two independent investigators.

*Western Blot Analysis*

Semi-quantitative immunoblot analysis were carried out in liver and kidney tissue samples as previously described [30]. Briefly, kidney and liver samples were homogenized in buffer and centrifuged (1320g, 5 mins, 4°C). To obtain the cytosolic fraction, supernatants were centrifuged (16125g, 4°C, 40 mins). The pelleted nucleolus were resuspended in extraction buffer and centrifuged (16125g, 20 mins, 4°C). Protein content was determined on both nuclear and cytosolic extracts using bicinchoninic acid (BCA) protein assay (Thermo Fisher Scientific Inc, Rockford, IL). Proteins were separated by 8% sodium dodecyl sulfate polyacrylamide (SDS-PAGE) gel electrophoresis and electrotransferred to polyvinylidene difluoride (PVDF) membrane. After blocking (1 hr in 10% dry milk solution), membranes were incubated with primary antibodies in 5% blocking solution overnight [rabbit anti-NF-κB (1:1000), rabbit anti-IKKα/β (1:1000), rabbit anti-Ser176/180 IKKα/β (1:5000), 1:5000 rabbit anti NLRP3 inflammasome (from Abcam), 1:1000 mouse anti-caspase 1 (p20) (from Adipogen)] followed by incubation with appropriated HRP-conjugated secondary antibodies. Proteins were detected with ECL detection system and quantified by densitometry using analytic software (Quantity-One; Bio-Rad, Hercules, CA). Results were normalized with respect to densitometric value of tubulin for cytosolic proteins or histone H3 for nuclear proteins.

### Supplemental Tables

#### Supplemental Table 1: Inclusion and exclusion criteria

| Including criteria | Excluding criteria |
|---|---|
| Blunt or penetrating trauma | Death in emergency room |
| ISS ≥ 16 | Death within 24 h of hospital admission |
| Age 18-80 | Known pre-existing immunological disorders |
| | Immunosuppressive medication |
| | Anti-coagulant medication |
| | Burns |
| | Con-comidant acute myocardial infarction |
| | Thromboembolic events |

#### Supplemental Table 2: Patients characteristics

| | **Trauma (n = 208)** |
|---|---|
| **Age (year) (IQR)** | 47.0 (31-60) |
| **Male sex (%)** | 156 (75.0) |
| **SOFA (points) (IQR)** | 5.00 (1.0-7.0) |
| **APACHE II (points) (IQR)** | 16.0 (6.0-22.0) |
| **ISS score (points) (IQR)** | 23.0 (17.0-32.0) |
| **LOS ICU (days) (IQR)** | 8.0 (4.0-15.0) |
| **LOS In-hospital (days) (IQR)** | 19.0 (13.0-29.0) |
| **MIF SR [pg/ml] (IQR)** | 6839 (3713-14205) |
| **MIF D2 [pg/ml] (IQR)** | 1598.0 (1080.0-2555.5) |
| **MIF D5 [pg/ml] (IQR)** | 1137.0 (650.0-1960.0) |
| **MIF D7 [pg/ml] (IQR)** | 1331.9 (814.2-2158.5) |

**Supplemental Figures**

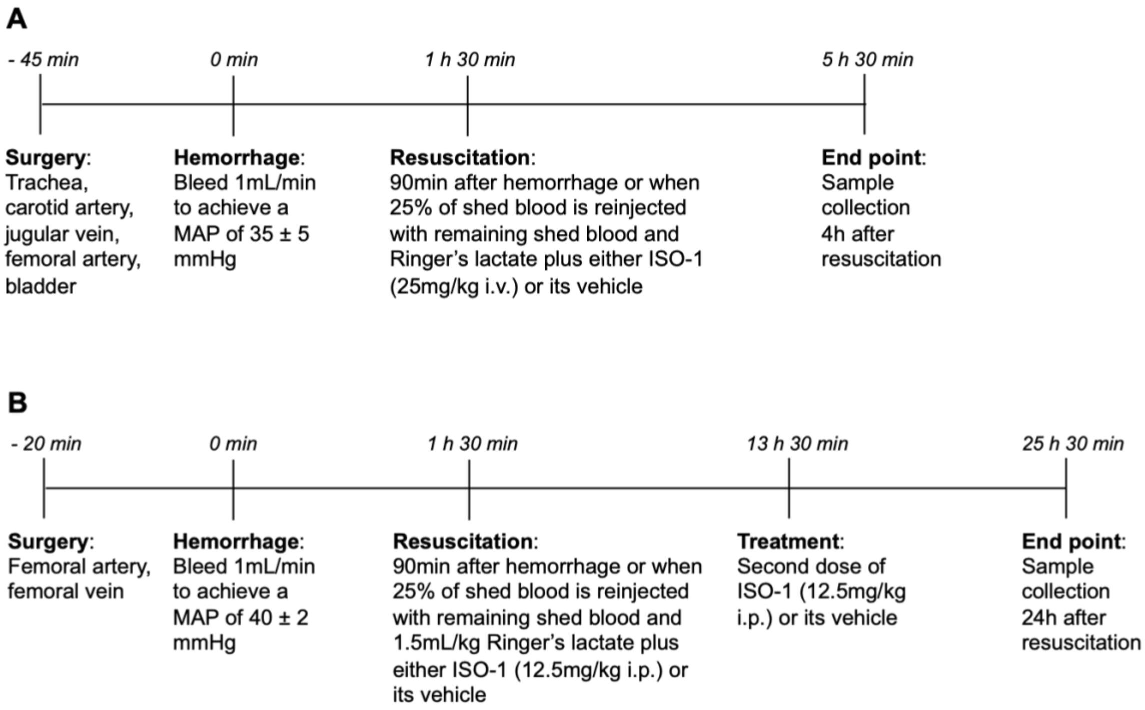


**Supplemental Figure 1: Schematic representation of the HS models.**

The experimental procedures at each stage of **(A)** the acute HS and **(B)** chronic model are shown.

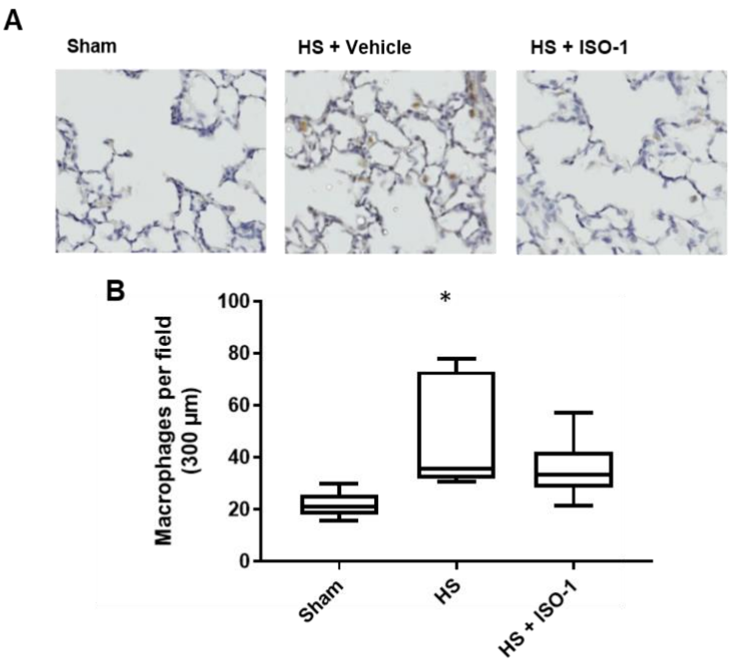


**Supplemental Figure 2: ISO-1 attenuates CD68+ cell infiltration in lungs after HS induction.**

(**A**) Representative images of immune stained lungs are presented. (**B**) CD68+ cell per filed in lungs in sham operated rats, after HS treated with vehicle and after HS treated with ISO-1. Data are expressed as box and whiskers blotted from min to max of 6-7 animals per group. Statistical analysis was performed using one-way ANOVA followed by a Bonferroni's post-hoc test.
